# Application of Generative Artificial Intelligence to Utilise Unstructured Clinical Data for Acceleration of Inflammatory Bowel Disease Research

**DOI:** 10.1101/2025.03.07.25323569

**Authors:** Alex Z Kadhim, Zachary Green, Iman Nazari, Jonathan Baker, Michael George, Ashley Heinson, Matt Stammers, Christopher M Kipps, R Mark Beattie, James J Ashton, Sarah Ennis

## Abstract

**Background:** Inflammatory bowel disease (IBD) research is a dynamic field. However, the growing volume of electronic health records (EHRs) and research data presents significant challenges. Traditional methods for structuring unstructured medical records are labour-intensive and lack scalability. Large language models (LLMs) may present a solution, yet their usefulness in data standardisation in the context of IBD remains unknown.

**Objective:** To evaluate the use of LLMs in structuring free-text histology and radiology reports from IBD patients, compare their performance to manual clinician curation, and assess the usefulness of fine-tuning and retrieval-augmented generation (RAG).

**Design:** We developed an IBD-specialised LLM-based framework utilising structured prompt engineering and fine-tuning. Reports were manually curated and processed using various LLMs. Performance was assessed and RAG was used to enhance model responses with clinical guidelines from European Crohn’s and Colitis Organisation (ECCO) and the European Society for Paediatric Gastroenterology Hepatology and Nutrition (ESPGHAN).

**Results:** Overall, Llama 3.3 achieved the highest F1 for histology and imaging (1 ± 0 and 0.85 ± 0.29, respectively) in extracting findings and anatomical regions, surpassing other models in structured data generation. Fine-tuning improved the performance of the smaller Llama 3.1 8B model for imaging reports (0.7 ± 0.46 vs 0.82 ± 0.35), enabling better extraction with reduced computational requirements.

**Conclusion:** Our findings demonstrate the feasibility of LLM-based automated structuring of IBD-related medical records. Unstructured data from free text reports can be reliably converted to standardised ontologies with location, severity, and qualifiers. These advancements enable scalable, privacy-compliant AI-driven solutions for data standardisation.

**VISUAL ABSTRACT:** 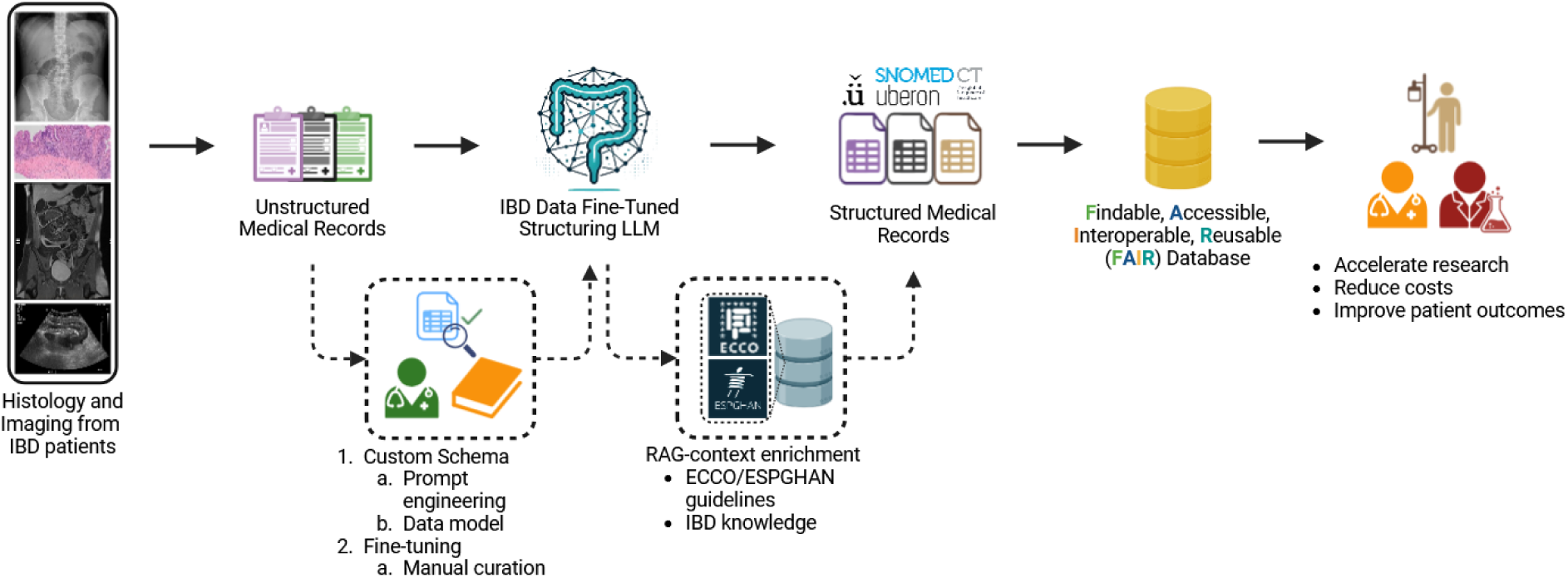

**Key Messages:** *What is already known on this topic:* Traditional methods for structuring unstructured medical records for research are labour-intensive and lack scalability. IBD patients generate vast quantities of longitudinal medical data due to the chronicity of disease. Large language models (LLMs) are well-positioned for data extraction and standardisation purposes.

*What this study adds:* This study demonstrates that Llama 3.3-70B and fine-tuned smaller models (Llama 3.1 8B) can accurately structure IBD-related histology and radiology reports. Additionally, retrieval-augmented generation (RAG) enhances clinical interpretability by incorporating guideline-based context.

*How this study might affect research, practice or policy:* The use of LLMs in structuring EHR data can significantly accelerate IBD research, improve data standardisation, and facilitate privacy-compliant AI-driven solutions for clinical decision support and policy development.

## INTRODUCTION

Inflammatory bowel disease (IBD) is a chronic, relapsing, and remitting group of disorders, traditionally categorised as Crohn’s disease (CD), ulcerative colitis (UC), and IBD-unclassified (IBDU) [1]. IBD research is dynamic, with a rapidly evolving therapeutic landscape, however, personalisation and prognostication remain limited [2]. The advent of big data and artificial intelligence (AI) methods has begun to improve personalised approaches [3,4]. Growing volumes of data give rise to new challenges for data integration and analysis, delaying actionable changes which can improve patient outcomes [5]. Specifically, disparate data types such as electronic health records (EHRs), imaging outputs, and high throughput ‘omics data present data management issues [6]. Reliable and trustworthy methods for converting rich free-text data to structured, standardised ontologies sources are needed to harness the value of these data for patient benefit [7,8].

FAIR data principles, which describe data as Findable, Accessible, Interoperable, and Reusable, provide guidance to improve data quality, accelerate medical research output, and, ultimately, positively impact patient outcomes [9]. At present, the lack of FAIR data limits data utilisation for research [10]. Large language models (LLMs) are a type of neural network specialised for text that have been successfully used in medical research for assisting in patient and clinician decisions, summarising medical reports, structuring unstructured data, and determining causal genes in genome wide association studies (GWAS) [11–16]. Several proprietary LLMs such as GPT-4 and Claude exist, but these models present data protection issues as sensitive patient data must be sent to hosted servers [17,18]. Conversely, open-source LLMs such as Llama, Mistral, Gemma, and others can be locally installed within secure data environments [19]. Table 1 presents a list of commonly used terminology summarising AI methods, ontologies, and software packages.

**Table 1:** Commonly used terminology.

|  | Term | Definition |
| --- | --- | --- |
| AI & Language Models | **Large Language Model (LLM)** | A type of artificial intelligence model designed to process and generate human-like text based on vast amounts of training data. |
|  | **Prompt Engineering** | The practice of designing and optimising input prompts to improve LLM performance and response accuracy. |
|  | **Fine-Tuning** | The process of adapting a pre-trained LLM to a specific task or domain using additional training data. |
|  | **Retrieval-Augmented Generation (RAG)** | A technique that enhances LLM outputs by retrieving relevant external data, such as clinical guidelines, before generating responses. |
|  | **Quantisation LoRA (QLoRA)** | A technique for reducing the memory requirements of fine-tuning LLMs by using low-rank adapters and quantised model weights. |
|  | **Supervised Fine-Tuning (SFT)** | A method of improving LLMs using labelled training data to guide the learning process. |
|  | **Semantic Embeddings** | A method of representing words, phrases, or documents as numerical vectors in a high-dimensional space, capturing their semantic meaning and relationships. |
| Ontologies | **SNOMED CT** | A comprehensive clinical terminology used for electronic health records (EHRs) and standardised medical documentation. |
|  | **Human Phenotype Ontology (HPO)** | A structured vocabulary describing phenotypic abnormalities associated with human diseases. |
|  | **National Cancer Institute Thesaurus (NCIT)** | A terminology system used primarily for cancer research, genomics, and drug development. |
|  | **Uberon Anatomy Ontology (UBERON)** | A cross-species anatomical ontology used to standardise anatomical structures. |
|  | **International Classification of Diseases (ICD-10)** | A globally used system for diagnosing and classifying diseases and conditions. |
|  | **Medical Dictionary for Regulatory Activities (MedDRA)** | A standardised medical terminology for adverse event classification in pharmacovigilance and clinical trials. |
|  | **Mondo Disease Ontology (MONDO)** | A disease classification system integrating multiple disease ontologies for harmonisation across biomedical databases. |
| Software Packages | **OntoGPT** | A Python package that enables structured extraction of medical and scientific information from free-text using LLMs. |
|  | **BioPortal Annotator** | A web-based tool that maps text to biomedical ontologies for interoperability and standardisation. |
|  | **Ollama** | A framework for running and managing LLMs locally, ensuring privacy and security for medical applications. |
|  | **FAISS** | Facebook AI Similarity Search, a tool for efficient nearest neighbour search used in retrieval-augmented generation. |
|  | **Unsloth** | A Python library designed for efficient fine-tuning of LLMs using techniques like QLoRA. |
|  | **Pteredactyl** | A free-text redaction tool that anonymises personally identifiable information (PII) in medical records. |

Low-rank adapters (LoRA) and quantisation LoRA (QLoRA) are methods of fine-tuning open-source LLMs that enable adaptation by significantly reducing memory requirements through quantising model weights, thus facilitating deployment in secure, resource-limited environments [20,21]. Retrieval augmented generation (RAG) utilises a retrieval mechanism with a LLM, enabling closed (or open-) source models without internet, the ability to enhance their responses by retrieving relevant information (e.g. clinical guidelines) from a local or curated database [22]. Therefore, fine-tuning and RAG methods are powerful enhancement techniques that improve the accessible deployment of specialised LLMs in secure data environments.

We aimed to develop an IBD-specialised LLM-based package which extracts, structures, and standardises data-rich free-text clinical records such as histology and radiology reports. As histology and imaging reports are essential for diagnosing and monitoring IBD but remain largely unstructured, extracting structured data from these reports enables large-scale analysis of disease patterns and treatment responses. Thus, our model is designed to transform unstructured text into structured, timestamped outputs aligned with widely used clinical ontologies, including ICD-10, SNOMEDCT, and HPO terms, which are of particular importance to link phenotypes to genes [23]. This process ensures that key clinical information—such as diagnosis, pathological findings, treatment history, and disease progression—can be easily integrated into research-ready datasets. We provide this package as a freely accessible resource for the research community, enabling standardised data extraction across sites, facilitating federated analyses, and ultimately advancing our understanding of IBD.

## MATERIALS AND METHODS

### Electronic Health Record Acquisition

We obtained histology and imaging reports from a cohort of 1,573 research consented IBD patients (REC 09/H0504/125) from the University Hospital Southampton. Patient records were redacted using Pteredactyl, a publicly available, clinical, free-text redaction software developed in-house [24]. We obtained a total of 32,041 records comprised of n = 7,181 (22.41%) histological procedures and n = 24,860 (77.59%) imaging procedures from a cohort of 1,573 research-consented IBD patients spanning 28 years (1996 to 2024; Figure 1A). Reports comprised 13 different imaging modalities, including ultrasound (21.02%), magnetic resonance (17.25%), X-Ray (14.58), computerised tomography (12.52%), and miscellaneous or legacy (pre-2001) reports uncoded for procedure (17.25%), among various other subtypes (Figure 1B). An additional set of imaging reports (n = 30) was also obtained through MIMIC-IV, a publicly available database sourced from the EHRs of the Beth Israel Deaconess Medical Center [25].

**Figure 1:**
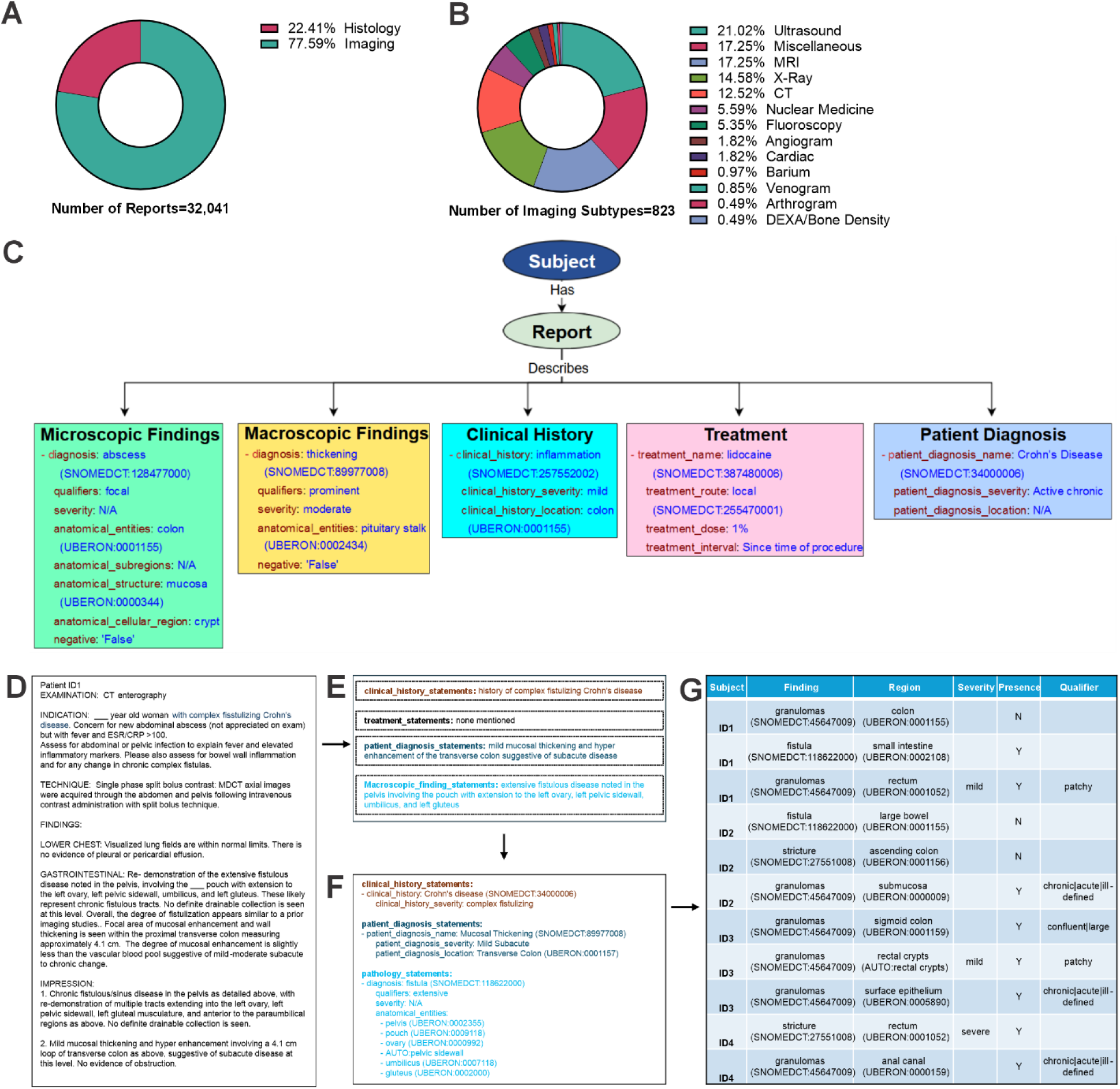
Findings from histology and imaging reports can be extracted and standardised into research ready outputs. **A)** Imaging and histology reports obtained from 1,573 IBD patients (n = 32,041). **B)** Breakdown of imaging subtypes found in patient reports (823 total subtypes). **C)** Semantic data model with seven entities (Subject, Report, Microscopic Findings, Macroscopic Findings, Clinical History, Treatment, and Patient Diagnosis) with example data input as obtained from patient records. OntoGPT-based workflow where **D)** an anonymised patient record is **E)** extracted into distinct semicolon-delimited statements and **F)** further standardised using ontological standards such as SNOMEDCT, NCIT, HPO, or UBERON for input into **G)** a FAIR database used for downstream analysis.

### Manual Curation

Manual curation of research-consented EHRs was performed by a clinical curation team composed of two paediatric gastroenterologists (ZG and JB). Due to the labour-intensive nature and time constraints, we randomly selected a subset of 90 reports from our local cohort, composed of an equal amount histology and radiology reports for independent curation (60 reports each with 30 overlapping). Curators were provided with a base schema which mirrored the LLM-provided schemas and written instructions for curating into the specified data model format, detailing clinical history, treatment, radiological (imaging) and microscopic (histology) findings (Figure 1C).

### Custom Data Model Creation

To contain extracted data and improve usability, a data model was developed to contain structured and standardised outputs from obtained imaging and histology reports (Figure 1C, Supplementary Figure 2). Each report was linked to a patient, given a unique identifier, and findings were categorised as pertaining to a patient’s clinical history, treatment, diagnosis, and macro- or microscopic findings. To account for the varying degree of granularity of anatomical locations described in different report types, we separated report findings by modality as histological (microscopic) and radiological (macroscopic) findings. This ensures standardised storage and retrieval of key patient findings, regardless of how the data was obtained.

### LLM-based Report Structuring and Iterative Prompt Engineering

We obtained several readily downloadable, open-source LLMs suitable for structuring data-rich medical records and adaptation. EHRs were structured and standardised using the iterative SPIRES method with the zero-shot learning-based OntoGPT v1.0.6 python package [26], which extracts statements and then standardises outputs. For a given report (Figure 1D), semicolon delimited statements matching our data model were first extracted as clinical history, treatment, patient diagnosis, and macro- or microscopic statements (Figure 1E). These statements were then matched to corresponding ontological standards for medical findings (SNOMEDCT, NCIT, or HPO) or anatomy (UBERON) for standardising outputs (Figure 1F). The resulting extracted and standardised findings are then able to be integrated into a tabular format (Figure 1G) with patient findings, region, severity, and qualifiers.

Performance was measured utilising a three-step approach where: 1) reports were structured in parallel by the team of gastroenterologists and all tested LLMs; 2) outputs were compared between each group, and, using iterative fine-tuning and schema generation; 3) the best performing model was further adapted (Figure 2). For initial comparisons, we tested the default OntoGPT package supplied pathology.yaml schema which broadly extracts details, providing minimal examples. We then progressed to develop two custom schemas: soton_IBD_histology.yaml (Supplementary Figure 1) and soton_IBD_imaging.yaml. To improve output results, prompts used within each schema were iteratively improved upon using manual curation results (e.g. integrating specific examples using manual curation results). LLMs were downloaded using Ollama v0.3.12 and interfaced within OntoGPT. All models were run using our high-performance compute (HPC) cluster IRIDIS X, on Dual NV-Linked A100 nodes (2x A100 per node, NV-Linked, 80 GB of VRAM per GPU). All models used and computational requirements are summarised in Table 2.

**Figure 2:**
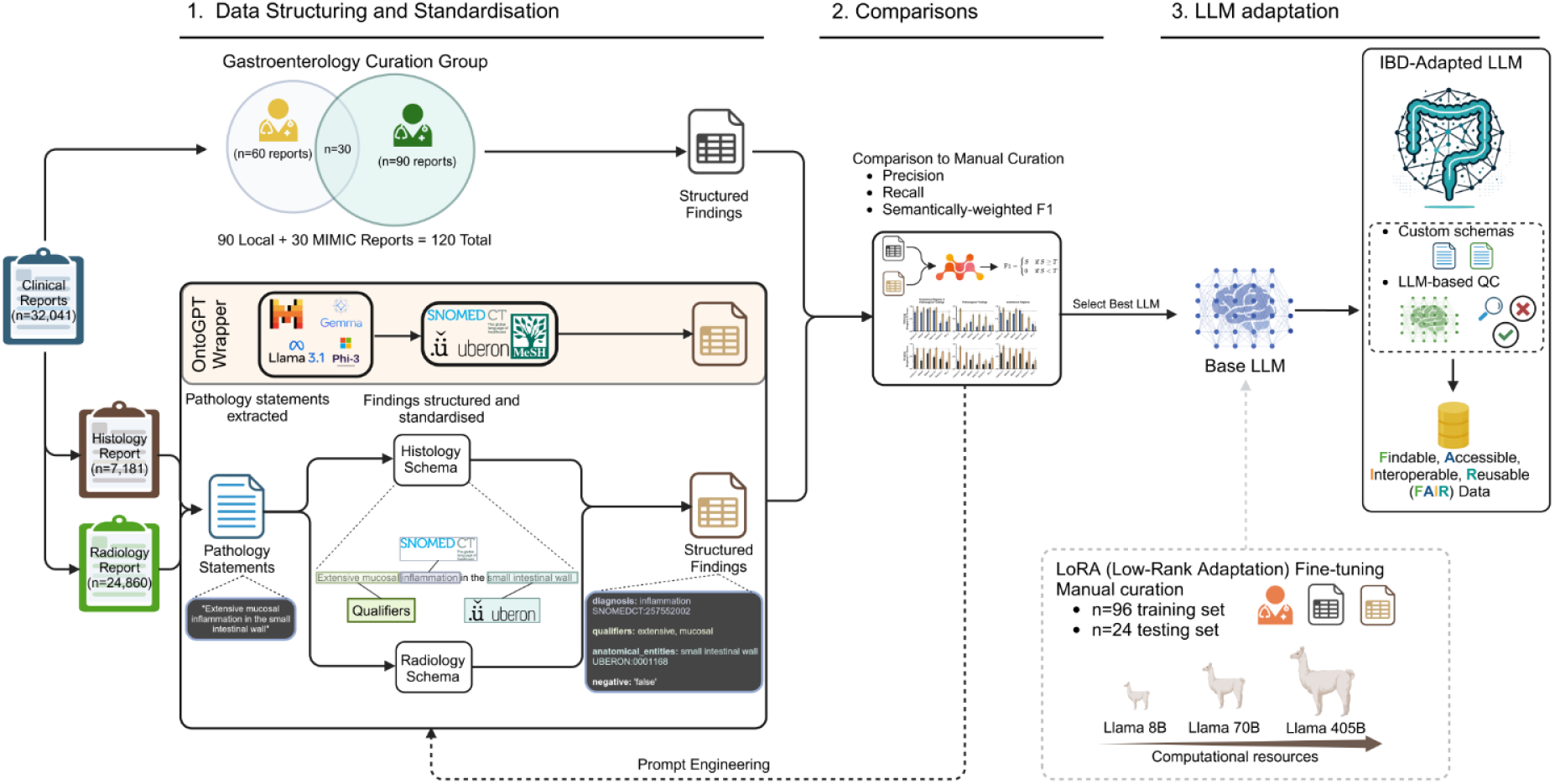
Schematic of IBD-adapted LLM creation and validation. *1. Data Structuring and Standardisation:* Histology (n = 7,181) and imaging (n = 24,860) reports were extracted and structured using OntoGPT in combination with several LLMs such as Llama 3, Gemma 2, Phi 3, and others. In parallel, a gastroenterology group manually curated a subset of reports (n = 90 local reports and n = 30 MIMIC external report; n = 120 total) with 30 overlapping reports to assess interrater variability. *2. Comparisons:* LLM structured findings were compared to manual curation using a semantically weighted scoring system to assess precision, recall, and variability of the finding and/or region meaning. In parallel, a custom schema was developed by iteratively incorporating prompt engineering from manual curation. *3. LLM adaptation:* the best performing model was selected, and low rank adaptation (LoRA) fine-tuning was used to input manually curated reports on models with varying sizes. Curated reports were split into a testing (n = 96) and training (n = 24) set for fine-tuning and validation, respectively. The IBD-adapted model was then tested, and LLM-assisted QC was used to produce high quality structured data.

**Table 2:** LLMs used in study.

| LLM | Size (GB) | Variation |
| --- | --- | --- |
| **mixtral** | 149 | mixtral:8x22b-instruct-v0.1-q8_0 |
| **llama3.1** | 149 | llama3.1:405b-instruct-q2_K |
| **llama3.3** | 141 | llama3.3:70b-instruct-fp16 |
| **llama3.1** | 141 | llama3.1:70b-instruct-fp16 |
| **mistral-large** | 130 | mistral-large:123b-instruct-2407-q8_0 |
| **qwen2.5** | 47 | qwen2.5:72b |
| **llama3.1** | 16 | llama3.1:8b-instruct-fp16 |
| **gemma2** | 5.2 | gemma2:2b-instruct-fp16 |
| **phi3** | 4.1 | phi3:3.8b-mini-4k-instruct-q8_0 |

### Comparisons to Manual Curation

To evaluate the alignment between manually curated data and outputs from LLMs, we computed similarity-based precision, recall, and F1 scores using semantic embeddings [27]. Data from manually structured reports and LLM-generated outputs were pre-processed by normalising text, removing numerical identifiers (e.g., SNOMED CT and UBERON codes), and aggregating structured information for each report. Sentence embeddings were generated using the mxbai-embed-large model via the Ollama application programming interface (API), with a caching mechanism to optimise computation. The caching was strictly limited to embeddings and was cleared between runs to prevent residual memory retention. Cosine similarity was used to quantify semantic overlap between manual and model outputs, and a threshold of 0.8 (acceptable match) was applied to determine matches [28]. We performed manual validation on a subset of cases to assess the accuracy of the similarity-based matching. If similarity exceeded this threshold, precision, recall, and F1 were assigned the similarity value; otherwise, they were set to zero. Aggregated results were stored and analysed to compare LLM outputs across multiple datasets.

### LLM Fine-Tuning

A Llama [29] 3.1-8B-Instruct model was fine-tuned to generate structured pathology statements from free-text histology reports using a parameter-efficient fine-tuning (PEFT) approach with LoRA [30]. Reports were separated into a testing (n = 24) and training (n = 96) set and converted into an input and completion format for training (323 total data points in total):

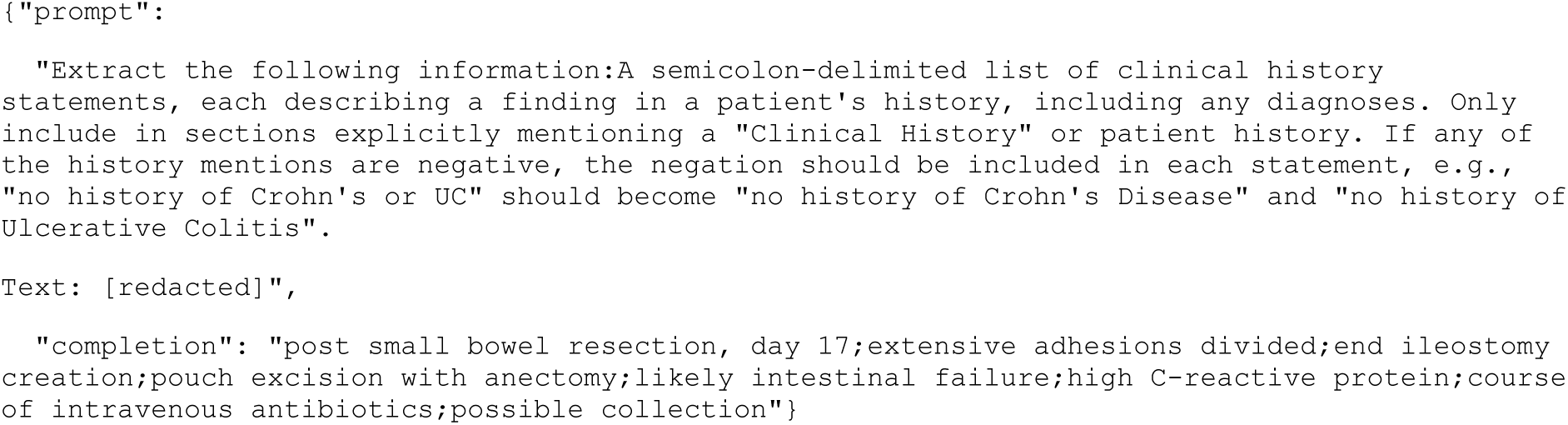

### BioPortal Annotator Standardisation

To facilitate ontology interconversion and standardisation of medical terminology, we utilised the BioPortal Annotator API [31]. Input terms were extracted from a TSV file and processed via API queries, retrieving ontology-aligned concepts, unique identifiers, and synonyms. The returned mappings were parsed to extract relevant class labels, concept identifiers, and alternative terminologies. The process prioritised longest match concepts to maximise specificity, and each term was cross-referenced across multiple ontologies for consistency.

### HPO Mapper

Mapping patient findings to Human Phenotype Ontology (HPO) [32] terms can be challenging due to the ontology’s structure, which often integrates both pathological findings and anatomical regions within a single term. To address this, a semantic similarity approach was employed using nomic-embed-text embeddings to match clinical findings and anatomical locations to the most relevant HPO term. Patient reports stored in JSON format were parsed to extract findings and their corresponding anatomical regions, which were then embedded and compared against a precomputed HPO embeddings database using cosine similarity. Matches exceeding a similarity threshold of 0.74, as defined by incremental adjustment and confirmation of adequate matches, were assigned as the most relevant HPO term. The final mapped results, including HPO IDs, term names, and subsequently mapped gene names were stored in structured CSV format for downstream clinical and research applications.

### LLM-assisted Final Quality Control and Flagging

A quality control (QC) pipeline was implemented to evaluate the accuracy of structured pathology extractions using a Llama 3.3-70B-Instruct model. Extracted objects from structured reports were parsed from YAML files, and each was assessed against the original input text for completeness and correctness. The QC process involved querying the LLM to identify missing, incorrect, or extraneous findings in the extracted data:

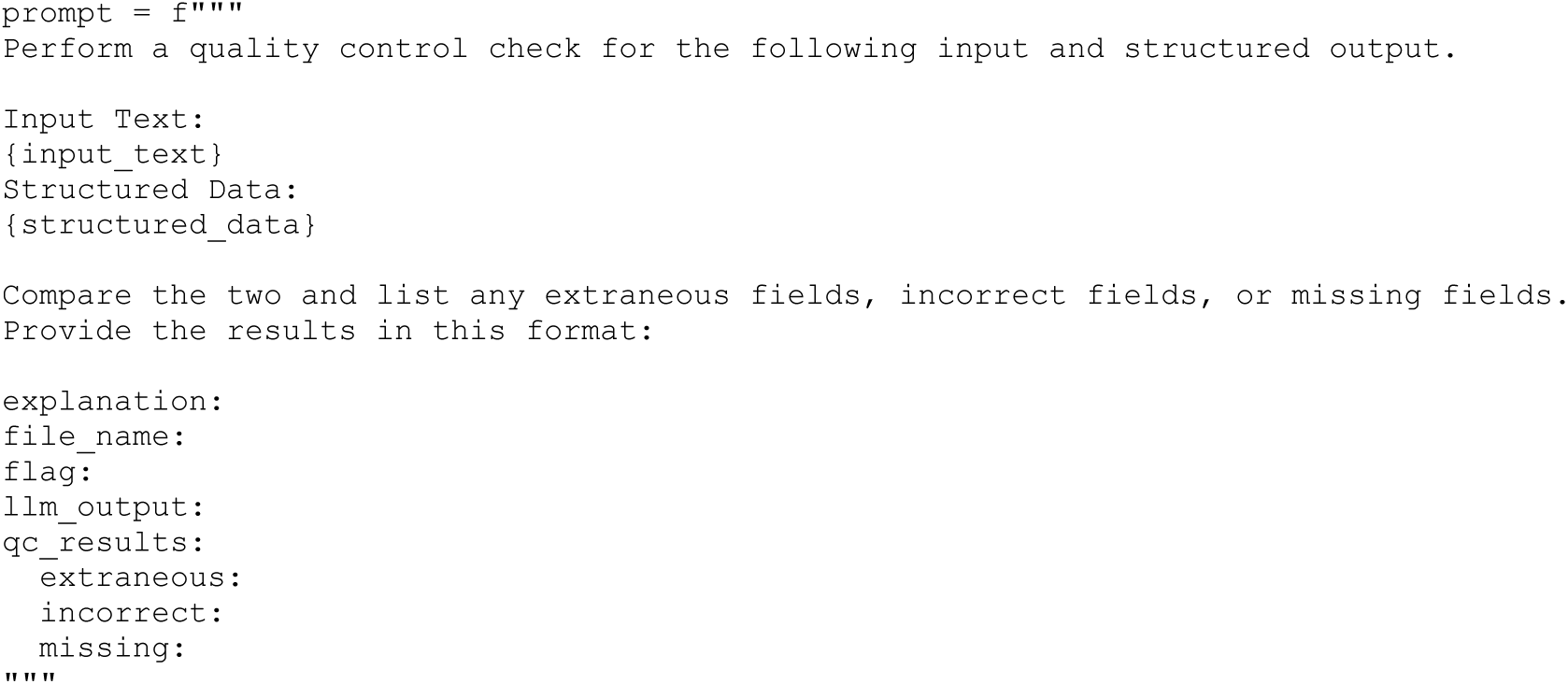

The model was prompted to compare the structured output against the raw text and return a structured response, flagging discrepancies. To ensure robustness, queries were executed with a context size of 16,000 tokens and retried up to three times in case of incomplete responses. The results were parsed to extract identified issues, and a binary flag was assigned to each report to indicate whether discrepancies were present.

### RAG-based output enrichment

A retrieval-augmented generation (RAG) pipeline was developed to enhance structured pathology outputs with relevant treatment recommendations using a Facebook AI Similarity Search (FAISS)-based semantic search. Embeddings for free-text pathology statements were generated using mxbai-embed-large and stored in a FAISS index [33] for efficient similarity retrieval. Semantic similarity search was performed against precomputed embeddings to retrieve the top five most relevant clinical context snippets. The retrieved context was then used to construct an enriched prompt incorporating relevant treatment guidelines, which the Llama 3.3-70B-Instruct model subsequently processed via the Ollama API to generate structured treatment recommendations. Each suggested treatment was aligned with the retrieved context and included source citations. The final enriched outputs were stored for downstream analysis.

## RESULTS

### LLMs can be used to extract findings from EHRs

Structuring IBD imaging and histology reports enhances their clinical utility for disease classification, prognosis, and research integration. Thus, manual curation of EHRs was performed using structured schemas (Supplementary Figure 1), a custom data model was developed for standardisation (Figure 1C), and LLMs were iteratively refined and evaluated against manual curation using semantic similarity-based metrics (Figure 2). We compared the semantic meaning of LLM and manually curated outputs by generating embeddings to convert words into mathematical representations of their meaning (Figure 3A). Interrater scoring between curators showed an average semantically weighted precision and recall (F1) of 0.9 ± 0.11 (Supplementary Figure 3), suggesting some variability among curators, as expected. We also compared a base extraction schema, (without specific prompt engineering or exemplar output to guide the model), and a prompt engineered schema.

**Figure 3:**
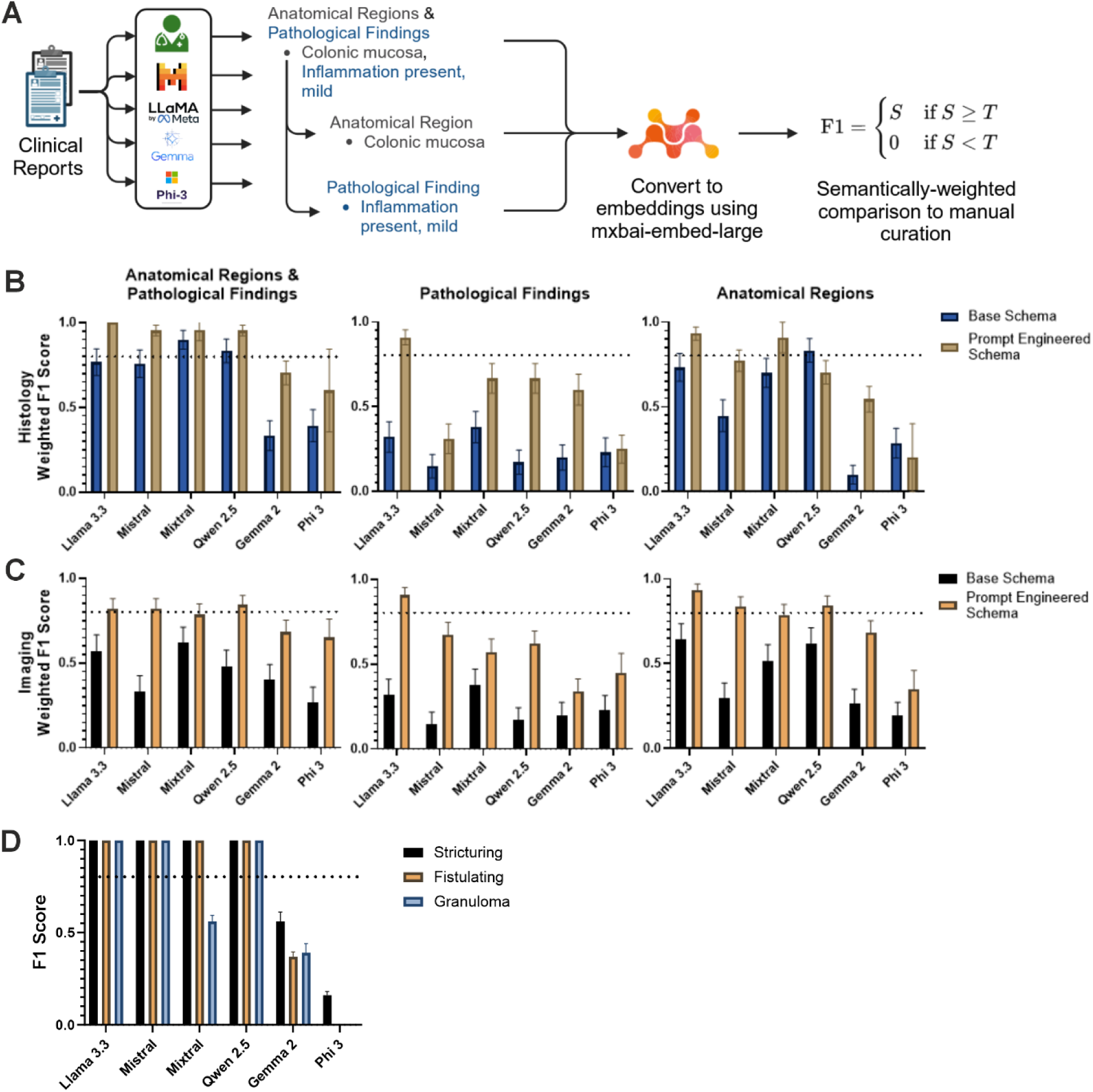
LLaMa 3.3 can extract patient findings and anatomical regions with high fidelity. **A)** Schematic of report comparison method where anatomical regions and pathological findings are compared together and separately to manual curation by conversion to embeddings and calculating semantically weighted F1 (0.8 similarity threshold). **B)** Results of LLM comparison between LLaMa 3.3, Mistral, Mixtral, Qwen 2.5, Gemma 2, and Phi 3 vs manual curation for histology reports (n = 60) and **C)** imaging reports (n = 60). **D)** Comparison between manual and LLM curation for extraction of structuring, fistulating, or granuloma from imaging and histology reports (n = 120). Data are represented as mean ± SEM.

Comparing Llama 3.3, Mistral, Mixtral, Qwen 2.5, Gemma 2, and Phi 3, we found that Llama 3.3 consistently scored above 0.8 for F1 across all report types (Figure 3B, Table 3). When combining anatomical regions with pathological findings for comparisons, all models showed improved performance with prompt engineering (Figure 3B, C; Table 3). For pathological findings in isolation, Llama 3.3 was the only model demonstrating acceptable performance (F1 > 0.8), whereas Llama 3.3, Mistral, and Mixtral performed well at extracting anatomical regions. Overall, the larger models tested (Llama 3.3, Mistral, Mixtral, and Qwen 2.5) performed better than smaller models with and without prompt engineering, as expected. When examining specific findings such as the presence of strictures, fistulas, or granulomas, most larger models tested were able to extract these with high precision and accuracy (Figure 3C). These findings demonstrate that Llama 3.3 can consistently extract anatomical and pathological findings from medical reports using our custom prompt-engineered schema. Moreover, extracting these disease features enables the generation of clinically and patient-relevant longitudinal phenotype data, which can be integrated into multi-omics research.

**Table 3:** F1 Scores for tested models.

|  |  | Anatomical Regions & Pathological Findings |  | Pathological Findings |  | Anatomical Regions |  |
| --- | --- | --- | --- | --- | --- | --- | --- |
| Histology | Llama 3.1 8B | 0.93 | 0.26 | 0.77 | 0.43 | 0.65 | 0.48 |
|  | Llama 3.1 70B | 0.98 | 0.15 | 0.98 | 0.15 | 0.80 | 0.41 |
|  | Llama 3.3 70B | 1.00 | 0.00 | 0.91 | 0.29 | 0.93 | 0.25 |
|  | Llama 3.1 405B | 0.98 | 0.15 | 0.70 | 0.46 | 0.84 | 0.37 |
|  | Llama 3.1 8B FT | 0.96 | 0.16 | 0.89 | 0.23 | 0.80 | 0.39 |
|  | Llama 3.3 70B FT | 1.00 | 0.00 | 0.91 | 0.29 | 0.94 | 0.16 |
|  | Mistral | 0.95 | 0.21 | 0.31 | 0.47 | 0.77 | 0.42 |
|  | Mixtral | 0.96 | 0.21 | 0.67 | 0.48 | 0.91 | 0.30 |
|  | Qwen 2.5 | 0.95 | 0.21 | 0.67 | 0.48 | 0.70 | 0.46 |
|  | Gemma 2 | 0.70 | 0.46 | 0.60 | 0.50 | 0.55 | 0.50 |
|  | Phi 3 | 0.60 | 0.55 | 0.25 | 0.44 | 0.20 | 0.45 |
| Imaging |  |  |  |  |  |  |  |
|  | Llama 3.1 8B | 0.70 | 0.46 | 0.64 | 0.49 | 0.73 | 0.45 |
|  | Llama 3.1 70B | 0.80 | 0.40 | 0.73 | 0.45 | 0.82 | 0.39 |
|  | Llama 3.3 70B | 0.82 | 0.39 | 0.91 | 0.29 | 0.93 | 0.25 |
|  | Llama 3.1 405B | 0.91 | 0.29 | 0.70 | 0.46 | 0.89 | 0.32 |
|  | Llama 3.1 8B FT | 0.82 | 0.35 | 0.80 | 0.32 | 0.81 | 0.39 |
|  | Llama 3.3 70B FT | 0.85 | 0.29 | 0.93 | 0.16 | 0.90 | 0.32 |
|  | Mistral | 0.82 | 0.39 | 0.67 | 0.47 | 0.84 | 0.37 |
|  | Mixtral | 0.79 | 0.42 | 0.57 | 0.50 | 0.79 | 0.42 |
|  | Qwen 2.5 | 0.84 | 0.37 | 0.62 | 0.49 | 0.84 | 0.37 |
|  | Gemma 2 | 0.68 | 0.47 | 0.34 | 0.48 | 0.68 | 0.47 |
|  | Phi 3 | 0.65 | 0.49 | 0.45 | 0.51 | 0.35 | 0.49 |

### Llama 3.3 70B outperforms Llama 3.1 405B in data standardisation

Since Llama 3.3 performed optimally in testing, we next sought to explore its variations to assess the performance of smaller and larger Llama 3 variations. Therefore, we compared variations of Llama 3.x, including Llama 3.1 8B, 70B, 405B, and Llama 3.3 70B using our prompt engineered schema (Figure 4A–D). Due to its heavy computational requirements, a quantised version (q2_K) of Llama 3.1 405B was used. Across all report types, both Llama 3.1 and 3.3 70B consistently scored highly (average F1 > 0.8), compared to the quantised 405B (F1 = 0.71 ± 0.46) when extracting pathological findings, due to loss of precision caused by quantisation. The smallest model, Llama 3.1 8B, demonstrated good performance with histology (F1 = 0.93 ± 0.26), but not imaging reports (F1 = 0.70 ± 0.46). The performance of all models improved when extracting findings and regions from histology reports compared to histological reports. Llama 3.3 70B performed best for histology (F1 = 1 ± 0) and imaging (0.82 ± 0.39) compared to other variations. Thus, although Llama 3.1 8B can accurately extract data from histological reports, it is not well suited for imaging reports.

**Figure 4:**
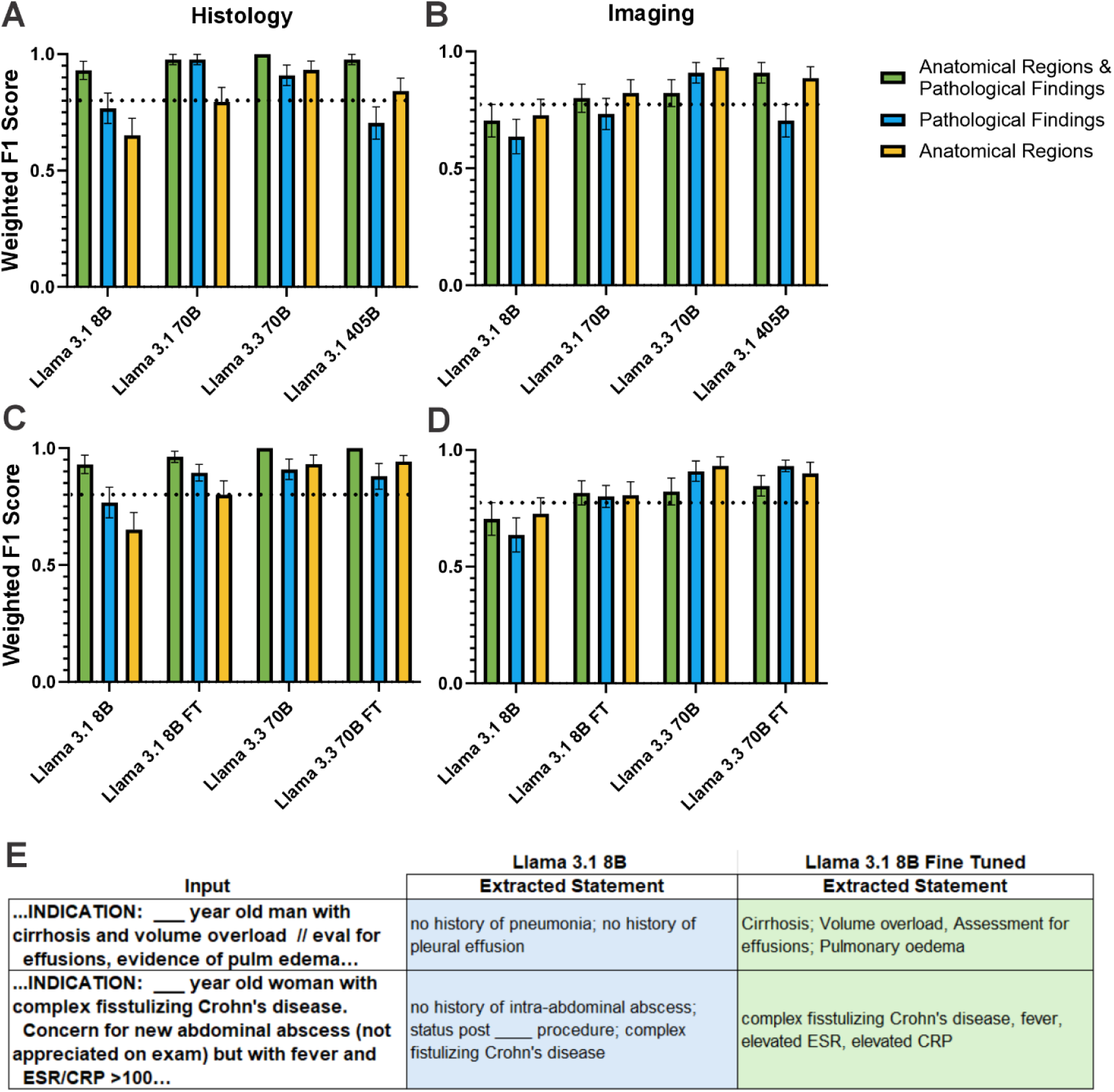
Fine-tuning LLaMa 3.1 8B with manual curation improves data extraction performance. **A)** Performance comparisons (semantically weighted F1 score) using prompt-engineered schema between variation of LLaMa 3.x with Llama 3.1 8B, 70B, 405B, and 3.3 70B for histology (n = 60) and **B)** imaging reports (n = 60). **C)** Performance comparisons between LoRA manual curation fine-tuned LLaMa 3.1 8B (Llama 3.1 8B FT) and fine-tuned LLaMa 3.3 70B (Llama 3.3 70B) to their respective base models for histology (n = 60) and **D)** imaging reports (n = 60). **E)** Representative table of extraction for a given imaging report between Llama 3.1 8B FT and base model. Data are represented as mean ± SEM.

### Fine-tuning Llama 3.1 8B improves data extraction performance

As larger LLMs have high computational requirements, we next sought to determine if the smaller Llama 3.1 8B model would benefit from fine-tuning using a set of manually curated reports. We first divided our reports into a training set (n = 96) and testing set (n = 24), representing 80% for training and 20% for testing. After fine-tuning, Llama 3.1 8B had an increased performance when structuring histology and imaging reports, comparable to Llama 3.1 70B (Figure 4C, D). No significant difference was observed between the base Llama 3.3 70B and fine-tuned model (Figure 4C, D). Examining the extracted terms, the fine-tuned Llama 3.1 8B model extracted more critical information from reports (Figure 4E).

These findings demonstrate that a fine-tuned Llama 3.1 8B can be used to extract and standardise data from EHRs efficiently.

### Extracted findings are interoperable between many ontologies

To facilitate ontology-based standardisation and conversions to other ontologies, we used the BioPortal Annotator API with built-in ontology mappings [31]. Standardised terms were converted across multiple biomedical ontologies, including HPO, NCIT, SNOMED CT, MEDDRA, MESH, DOID, MONDO, and ICD-10 (Figure 5A). While many clinical findings successfully matched existing ontology terms, HPO terms were frequently absent, particularly for histological findings encompassing both a pathological feature and an anatomical region. As HPO terms also provide mappings to genes, which are critical for downstream analysis relating phenotypes to genotypes [23], this limitation necessitated additional approaches to ensure comprehensive coverage of IBD-related HPO terms.

**Figure 5:**
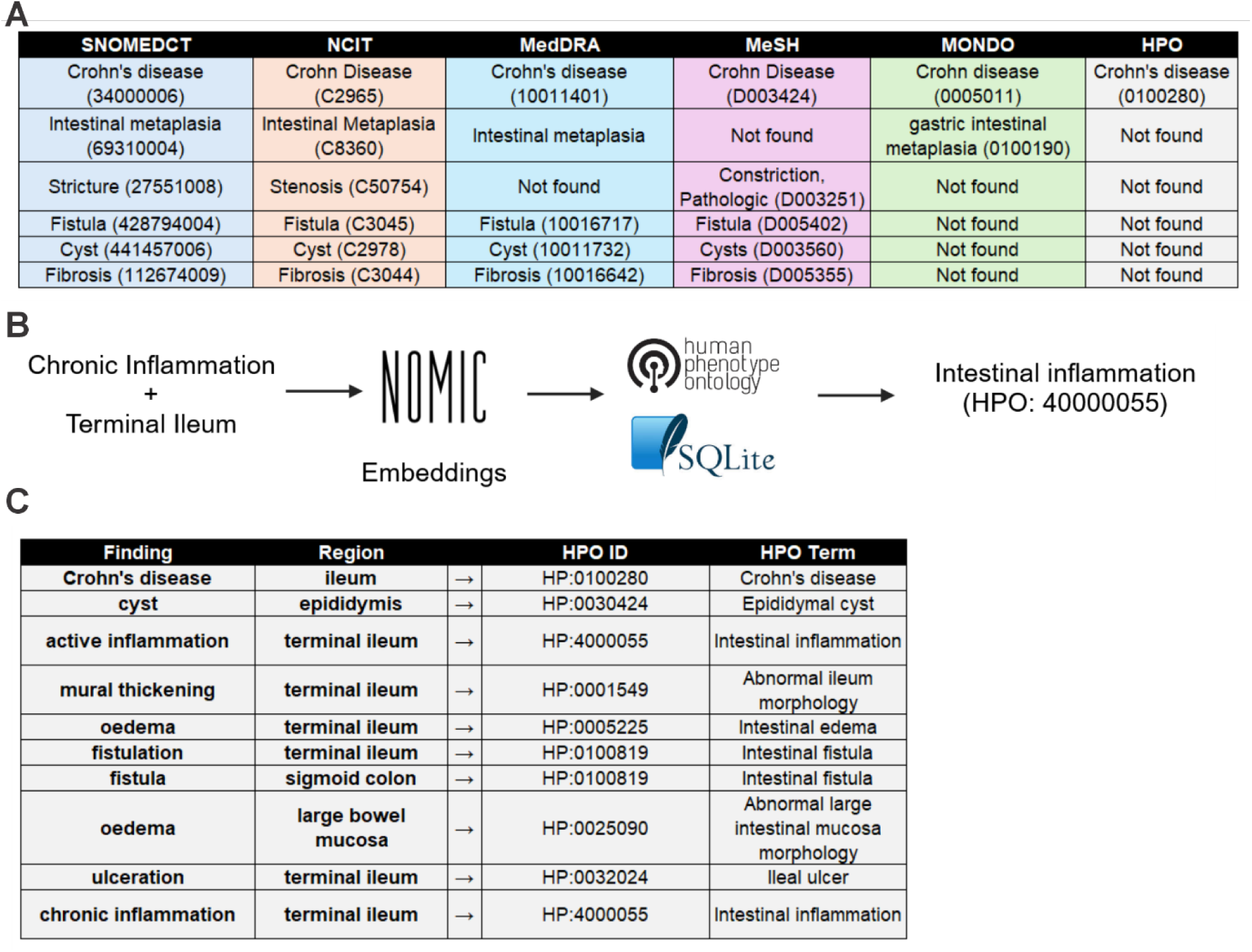
LLM-extracted findings are interoperable between several ontological standards. **A)** BioPortal annotator API output for mappings between terms in SNOMEDCT, NCIT, MedDRA, MeSH, MONDO, and HPO. **B)** Pipeline for RAG-based HPO annotation using combined findings and regions with **C)** representative output of HPO matching pipeline integrating findings and regions.

### RAG can be used to standardise to missed HPO terms

We implemented a RAG-based approach using semantic embeddings to address observed gaps to identify the most relevant HPO terms for patient findings (Figure 5B). We improved the identification of relevant HPO annotations, particularly for complex histological descriptions such as “oedema” (finding) + “terminal ileum” (region), which was converted to “intestinal edema” (HPO:0005225) (Figure 5C). Therefore, RAG allows for additional matching to HPO terms, often missed using the BioPortal Annotator and possibly other named-entity recognition techniques.

### LLM-assisted quality control can flag erroneous data extraction

Earlier sections detailed the iterative validation steps comparing LLM-generated results with human curation, reinforcing confidence in the structured data. Before formal QC is applied, additional checks are necessary to ensure robustness. Thus, we implemented a QC pipeline that systematically assessed extracted findings against the original free-text reports to evaluate the accuracy and consistency of LLM-generated structured reports. The QC system flagged discrepancies in three main categories: missing findings, incorrect findings, and extraneous findings (Figure 6A). Using the Llama 3.3-70B-Instruct model, reports were programmatically reviewed for inconsistencies, with structured prompts guiding the LLM to identify deviations from the original text. The QC checker successfully identified problematic reports where key pathology findings were either absent, misclassified, or included erroneously (Figure 6A, Supplementary Figure 4). Therefore, reports containing high levels of discrepancies are flagged for manual review, enabling curators to assess final outputs before integration into a FAIR database. This approach provided an automated validation layer, ensuring structured outputs aligned with clinical expectations while highlighting cases requiring further correction.

**Figure 6:**
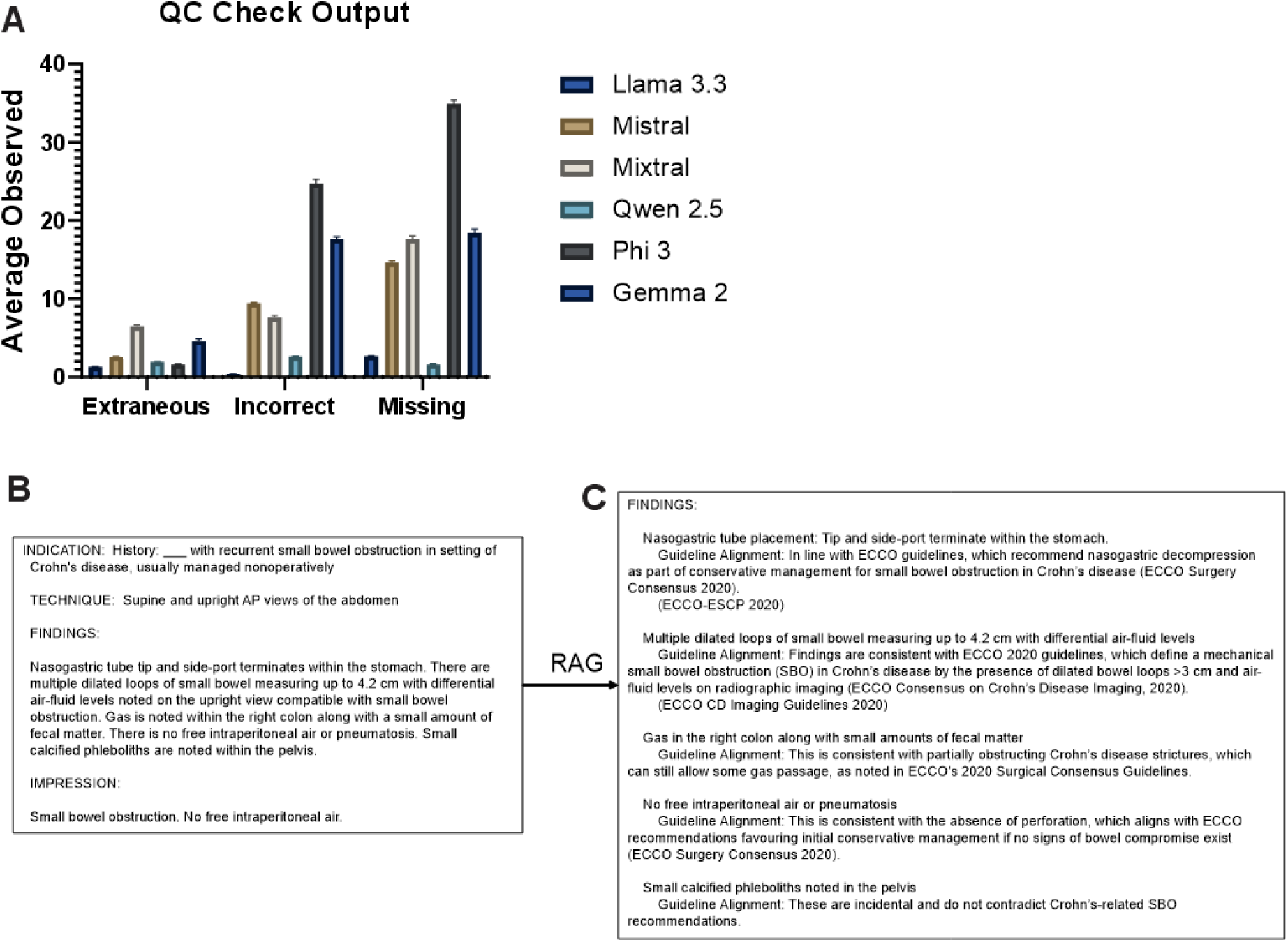
LLM-assisted QC checks and IBD guidelines can improve data quality and richness. **A)** Results from LLM-assisted QC flagging for reports curated by Llama 3.3, Mistral, Mixtral, Qwen 2.5, Phi 3, and Gemma 2 for histology and imaging reports (n = 120) showing extraneous, incorrect, or missing extracted findings per model. **B)** Imaging report after **C)** enrichment using RAG-based IBD (European Crohn’s and Colitis Organisation [ECCO] and the European Society for Paediatric Gastroenterology Hepatology and Nutrition [ESPGHAN]) guideline enrichment.

### RAG can be used to enrich findings with ECCO and ESPGHAN guidelines

To integrate evidence-based clinical guidance into structured pathology reports, we implemented a retrieval-augmented generation (RAG) approach that allows the LLM to retrieve and incorporate European Crohn’s and Colitis Organisation (ECCO) and the European Society for Paediatric Gastroenterology Hepatology and Nutrition (ESPGHAN) clinical guidelines [34–37] within its outputs. This method enables the model to contextualise extracted pathology findings, linking them to established diagnosis, treatment, and disease management recommendations. The model first processes the structured pathology report (Figure 6B) and retrieves the most relevant guideline sections using semantic search and embeddings. Pathology findings are supplemented with guideline-backed comments, enabling clinicians to interpret structured data within a clinically relevant framework (Figure 6C). This demonstrates the utility of RAG-based approaches for IBD research.

## DISCUSSION

Our findings demonstrate that LLMs can effectively extract and standardise unstructured clinical data from EHRs. In the future, this can provide a scalable, reproducible, and FAIR method for structuring histology and radiology reports for IBD research and clinical practice. LLM-based extraction can facilitate large-scale, reproducible analyses and enhance data harmonisation across institutions. Further, standardised outputs allow seamless integration with complementary datasets, including laboratory findings, genomics, and multi-omics profiles, enabling comprehensive disease characterisation and precision medicine approaches [5,38–40]. Such standardisation efforts are particularly critical in IBD research, where histological and radiological features, when linked with genetic and molecular data, can drive novel insights into disease pathophysiology, progression, and therapeutic response [1,2].

As we observed, larger parameter models such as Llama 3.3 70B consistently outperformed smaller counterparts due to improved complex reasoning [41]. This aligns with previous literature demonstrating the benefits of increased model size in medical natural language processing tasks, where more significant parameter counts allow for improved contextual understanding and generalisation [42]. On average, Llama 3 variants performed best when compared to similarly sized models. Although performance was better for the larger models tested, the computational requirements present challenges [43]. Our study using Llama 3.3 with two A100s observed an average processing time of approximately 39.46 seconds per report. For 32,041 reports, this would translate to an estimated 1,264,206 seconds (∼351.17 hours, or ∼14.6 days) of processing time under similar conditions. This problem could be exacerbated with the recent advent of newer models [44]. Overcoming these barriers while maintaining generalisability should be a primary focus for future research.

We found that fine-tuning smaller models, such as Llama 3.1 8B, with manually curated reports significantly improved performance, particularly for histology report structuring. This reduced the need for more computationally heavy LLMs and suggests that domain-specific adaptation remains a crucial strategy for optimising model performance in specialised medical tasks [45]. Additionally, smaller fine-tuned models can outperform larger base models [43]. Although fine-tuning requires high-quality labelled datasets, which are often labour-intensive, these efforts may be warranted to reduce time and computational resources necessary.

The inclusion of data from two distinct sources—our local patient cohort and the publicly available MIMIC-IV dataset [25]—allowed us to begin to test the generalisability of our approach across different institutional settings. Although studies have shown increased performance with small training sets and as few as 60 data points [46,47], expanding the dataset to additional sites and diverse healthcare systems would be beneficial to further assess model robustness and ensure the reliability of findings across varied clinical practices. Additionally, we have not yet examined the role of bias and propose this as a research tool. Furthermore, while our model demonstrated robust performance in structuring histology and imaging reports, additional validation on other EHR components would be valuable to assess broader applicability. We envisage this model will provide improved utilisation for patient benefit of large-scale molecular data generated through initiatives such as the IBD Bioresource and the International IBD genetics consortium [48,49].

In conclusion, our work highlights the potential of open-source LLMs for structuring unstructured EHRs in IBD, while demonstrating the importance of model size, fine-tuning, and RAG. Addressing current limitations through expanded multi-institutional validation, privacy-preserving comparisons with proprietary models, and further optimisation of computational efficiency will be critical next steps in advancing LLM-driven clinical data extraction. Integrating FAIR data principles into these workflows will further support reproducibility and accessibility, ultimately accelerating medical research and improving patient outcomes.

## Data Availability

Due to patient privacy restrictions, the underlying clinical data or fine-tuned models cannot be publicly shared. The fully fine-tuned models on 90 histology and imaging reports are available upon request.

https://github.com/UoS-HGIG/

https://huggingface.co/UoS-HGIG

## Data and Model Availability

Due to patient privacy restrictions, the underlying clinical data or fine-tuned models cannot be publicly shared. Therefore, we are providing Llama 3.1 8B adapters from 30 curated MIMIC-IV imaging reports [25], and all associated files for model deployment, available here: https://github.com/UoS-HGIG/ and https://huggingface.co/UoS-HGIG. The fully fine-tuned models on 90 histology and imaging reports are available upon request.

## Acknowledgements

The authors acknowledge the use of the IRIDIS High Performance Computing Facility, and associated support services at the University of Southampton in the completion of this work as well as the SETT data and AI team and CIRU. Above all, we would like to thank the patients and their families.

