## Supplementary material for "Application of Generative Artificial Intelligence to Utilise Unstructured Clinical Data for Acceleration of Inflammatory Bowel Disease Research": Suplementary Figures

### Supplementary Figures:

```yaml
id: http://w3id.org/ontogpt/soton_ibd_histology
name: soton_ibd_histology
title: University of Southampton IBD Histology Report Schema
description: >-
  A custom template for extracting and grounding histology reports focused on IBD pathologies.
license: https://creativecommons.org/publicdomain/zero/1.0/
prefixes:
  soton_ibd_histology: http://w3id.org/ontogpt/soton_ibd_histology/
  rdf: http://www.w3.org/1999/02/22-rdf-syntax-ns#
  PATO: http://purl.obolibrary.org/obo/PATO_
  UBERON: http://purl.obolibrary.org/obo/UBERON_
  SNOMEDCT: http://purl.bioontology.org/ontology/SNOMEDCT/
  ICD10CM: http://purl.bioontology.org/ontology/ICD10CM/
  NCIT: http://purl.obolibrary.org/obo/NCIT_
  MESH: http://purl.bioontology.org/ontology/MESH/
  MONDO: http://purl.obolibrary.org/obo/MONDO_
  linkml: https://w3id.org/linkml/
  pathology: http://w3id.org/ontogpt/soton_ibd_histology
keywords:
  - pathology
  - phenotype

default_prefix: soton_ibd_histology
default_range: string

imports:
  - linkml:types
  - core

classes:

  PathologyReport:
    tree_root: true
    description: >-
      A semicolon-delimited list of statements, each describing a pathology,
      including any diagnoses, one or more specific qualities being measured
      and the anatomical location or tissue the pathology is measured in. Each
      statement should describe a single condition.
    attributes:
      pathology_statements:
        description: >-
          A semicolon-delimited list of pathology statements, each describing a
          pathology, including any diagnoses, one or more specific qualities
          being measured and the anatomical location or tissue the pathology is
          measured in. If any of the pathology statements are negative, the
          negation should be included in each statment, e.g., "no granulomas or
          viropathic changes" should become "no granulomas" and "no viropathic
          changes".
        range: PathologyStatement
        multivalued: true
      clinical_history_statements:
        description: >-
          A semicolon-delimited list of clinical history statements, each
          describing a finding in a patient's history, including any diagnoses.
          Only include in sections explicitely mentioning a "Clinical History" or patient history.
          If any of the history mentions are negative, the negation should be included in each statment, e.g.,
          "no history of Crohn's or UC" should become "no history of
          Crohn's Disease" and "no history of Ulcerative Colitis".
        range: PatientClinicalHistory
        multivalued: true
      treatment_statements:
```

**Supplementary Figure 1: Soton IBD histology custom schema example.** Custom IBD-specific histology schema YAML input file used for OntoGPT based data structuring. Classes are defined which each description being used as a prompt for the LLM.

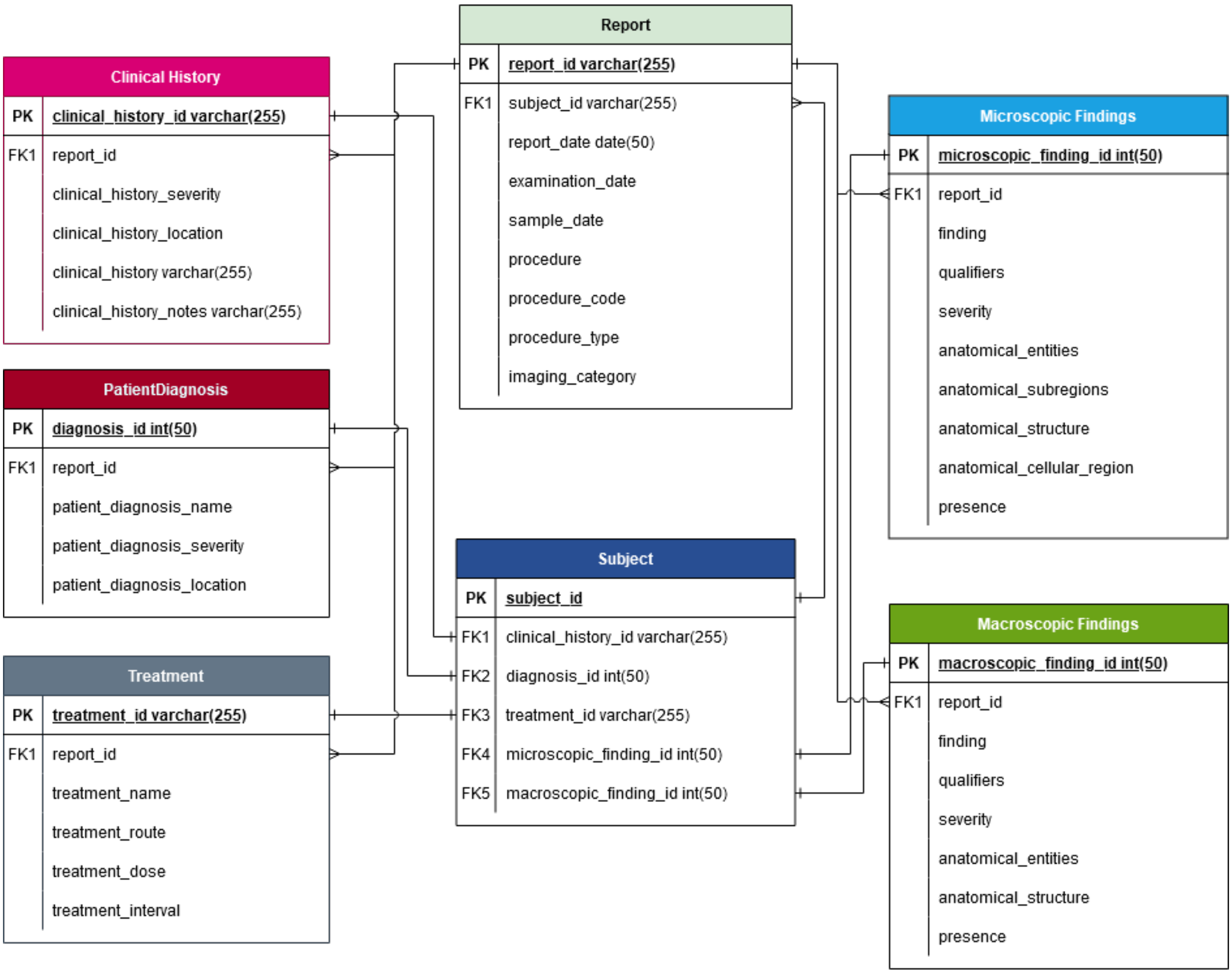


**Supplementary Figure 2: Complete physical data model.** Graphical representation of physical data model (relational database) containing seven entities: Report, Subject, Clinical History, Patient Diagnosis, Treatment, Microscopic Findings, and Macroscopic Findings with each primary key (PK) and foreign key (FK).

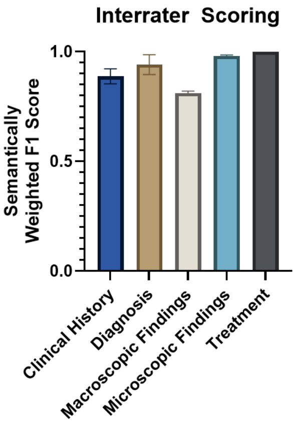


**Supplementary Figure 3: Interrater variability assessment.** Semantically weighted F1 score between manual curation by ZG and JB for extracted clinical history, diagnosis, treatment, and macro- and microscopic findings (n = 30). Data are represented as mean ± SEM.


```
explanation: '** The raw completion output closely matches the findings section of
  the input text, capturing all major points such as the presence of chain sutures/surgical
  clips, the non-obstructive bowel gas pattern, the absence of large air-fluid levels
  and free air, clear lung bases, and no evidence of bowel obstruction. The primary
  difference lies in the level of detail provided regarding the examination process
  itself (e.g., the obtaining of supine and upright views), which is not directly
  listed as a finding in the raw completion output but is integral to how the findings
  were obtained. Overall, the raw completion output accurately represents the key
  clinical findings from the input text without introducing inconsistent or unmentioned
  information.'
file_name: 16093709_16093709-RR-27_processed_macroscopic_findings.txt
flag: Y
llm_output: "Here is the comparison of the raw completion output with the input text:\n\
  \n* **Missing Findings:** \n  + The fact that supine and upright views of the abdomen\
  \ were obtained is not explicitly mentioned in the raw completion output, although\
  \ it is implied by the mention of various findings. However, the specific detail\
  \ about the views being obtained is not directly stated.\n\n* **Incorrect Findings:**\
  \ \n  + None identified. All findings in the raw completion output are consistent\
  \ with the input text.\n\n* **Extraneous Findings:** \n  + None identified. All\
  \ findings mentioned in the raw completion output are present in the input text.\n\
  \n* **Explanation:** The raw completion output closely matches the findings section\
  \ of the input text, capturing all major points such as the presence of chain sutures/surgical\
  \ clips, the non-obstructive bowel gas pattern, the absence of large air-fluid levels\
  \ and free air, clear lung bases, and no evidence of bowel obstruction. The primary\
  \ difference lies in the level of detail provided regarding the examination process\
  \ itself (e.g., the obtaining of supine and upright views), which is not directly\
  \ listed as a finding in the raw completion output but is integral to how the findings\
  \ were obtained. Overall, the raw completion output accurately represents the key\
  \ clinical findings from the input text without introducing inconsistent or unmentioned\
  \ information."
qc_results:
  extraneous:
  - '** '
  - '  + None identified. All findings mentioned in the raw completion output are
    present in the input text.'
  - '* **'
  incorrect:
  - '** '
  - '  + None identified. All findings in the raw completion output are consistent
    with the input text.'
  - '* **'
  missing:
  - '** '
  - '  + The fact that supine and upright views of the abdomen were obtained is not
    explicitly mentioned in the raw completion output, although it is implied by the
    mention of various findings. However, the specific detail about the views being
    obtained is not directly stated.'
  - '* **'
```


**Supplementary Figure 4: LLM-assisted QC output example.** Example output format of LLM-based QC flagging. Specific sections such as explanation, file_name, flag, llm_output, and qc_results allow further investigation into flagged reports by curators.
